# Methods and Operational Framework of the GALFLU Pragmatic Trial: Design and Feasibility of an Individually Randomized Controlled Trial Evaluating High-Dose Versus Standard-Dose Influenza Vaccination in Older Adults

**DOI:** 10.1101/2025.07.24.25332041

**Authors:** Narmeen Mallah, Jacobo Pardo-Seco, Carmen Rodriguez-Tenreiro-Sánchez, Iago Giné-Vázquez, Susana Mirás-Carballal, Marta Piñeiro-Sotelo, Martín Cribeiro-González, Mónica Conde-Pájaro, Juan-Manuel González-Pérez, Josefina Lorena Razzini, Irene Rivero-Calle, Ana Isabel Dacosta-Urbieta, Rebecca C Harris, Matthew M Loiacono, Robertus van Aalst, Joan Manel Farre, Marine Dufournet, Niklas Dyrby Johansen, Daniel Modin, Tor Biering-Sørensen, Carmen Duran-Parrondo, Federico Martinón-Torres, GALFLU trial team

## Abstract

**Background:** Optimizing influenza vaccination in older adults is a public health priority. While high-dose inactivated influenza vaccines (HD-IIV) have shown superior immunogenicity and efficacy over standard-dose vaccines (SD-IIV), there is a shortage of large-scale, individually randomized data evaluating real-world effectiveness against severe outcomes—especially within publicly funded health systems.

**Methods:** GALFLU is a pragmatic, individually randomized, open-label clinical trial comparing HD-IIV and SD-IIV in adults aged 65–79 years in Galicia, Spain, during the 2023–2024 and 2024–2025 influenza seasons. The trial embedded individual randomization into the regional vaccination program and exploited digital health registries for comprehensive, low-burden data collection under real-world conditions. The primary outcome is relative vaccine effectiveness (rVE) against a composite of influenza- or pneumonia-related hospitalizations. Secondary and exploratory outcomes include rVE against cardio-respiratory disease, laboratory-confirmed influenza, all-cause hospitalization, mortality, and healthcare utilization.

**Results:** GALFLU successfully demonstrated the feasibility of large-scale, pragmatic randomized controlled trials within routine clinical practice. Across both seasons, 133,822 participants were randomized (59,490 in 2023/24 and 74,986 in 2024/25), typically within 4–5 weeks per season. Automated linkage ensured >99% follow-up using electronic clinical and laboratory registries.

**Conclusions:** GALFLU provides an effective, scalable operational framework for generating robust real-world evidence through pragmatic, individually randomized trials embedded in public health systems. This model serves as a blueprint for future vaccine evaluations, supports regulatory and policy decisions with high-quality, population-level data, and illustrates the evolution toward “learning health systems” that integrate pragmatic trials within routine care.

## Introduction

Influenza continues to represent a significant global public health concern, with the World Health Organization (WHO) estimating up to one billion cases and 3– 5 million severe episodes each year.^1^ Vulnerable groups—including the elderly, individuals with comorbidities, young children, and the immunocompromised— face the greatest risks of complications and mortality.^2,3^ Among individuals aged 75 years and older, influenza is estimated to contribute to between 51.3 and 99.4 annual deaths per 100,000.^1,4^

Despite widespread vaccination efforts, the effectiveness of standard-dose inactivated influenza vaccines (SD-IIV) is notably lower in older compared to younger adults (30.6% versus 36.7%), highlighting the need for alternative vaccination strategies in this population.^5^ High-dose inactivated influenza vaccine (HD-IIV) has demonstrated superior protection against laboratory-confirmed influenza in randomized trials,^6^ and meta-analyses of observational data and smaller RCTs suggest HD-IIV may also reduce influenza-related, pneumonia-related, and all-cause hospitalizations, as well as respiratory and cardiovascular outcomes.^7,8^

Consequently, health authorities have increasingly recommended HD-IIV for older adults.^9,10^ In Galicia, Spain, the quadrivalent HD influenza vaccine (QIV-HD; Efluelda®) has been available since April 2020 for individuals aged 65 and older and is currently recommended for those aged ≥80 or for nursing home residents aged ≥60, per the 2023/2024 local policy.^11^ There is significant public health interest in understanding the potential impact of HD-IIV versus SD-IIV on clinically relevant endpoints, such as influenza- or pneumonia-related hospitalizations, particularly in the 65–79 age group. Traditional explanatory RCTs, while offering insights into efficacy under ideal conditions, often fail to reflect heterogeneity in the general population and the organizational realities of vaccination under routine clinical conditions. As a result, their findings may have limited generalizability or external validity for population-level decision-making.^12^

Pragmatic trial designs are specifically intended to address these limitations by evaluating interventions in the context of routine care. Embedding randomization in large-scale, real-world vaccination programs—and relying on population-based health registries—preserves the rigor of RCTs while maximizing applicability and efficiency. Individual randomization remains the gold standard for causal inference, minimizing confounding by evenly distributing known and unknown risk factors. However, robust comparative effectiveness evidence for HD-IIV from individually randomized pragmatic trials remains scarce. The recent feasibility study in Denmark (DANFLU-1) assessed clinical endpoints in an exploratory fashion but was not powered to detect meaningful differences.^13^ DANFLU-2 is an ongoing trial but alone cannot deliver on globally generalizable results.

To address this evidence gap, the GALFLU trial was initiated as a pragmatic, registry-based, open-label, active-controlled, individually randomized study in Galicia, Spain, to assess the real-world effectiveness of HD-IIV versus SD-IIV in adults aged 65–79 years.

The availability of comprehensive, population-based health registries in Galicia enables the efficient implementation and follow-up of large-scale pragmatic trials. By capturing endpoint and resource use data passively—without the need for medical record abstraction or additional patient follow-up—such registries support the detection of both common and rare clinical endpoints, increasing statistical power while reducing cost and burden. Findings from GALFLU can contribute to cost-effectiveness analyses and inform future policy for influenza vaccination in older adults.

### Objective of this paper

This article describes the rationale, design, operational framework, and data sources of the GALFLU trial^14^. Study outcomes and analyses will be reported in subsequent publications; this methods paper focuses on trial design and implementation.

## Methods

### Setting and Population

GALFLU was conducted in Galicia, an autonomous region in northwestern Spain with nearly 2.7 million inhabitants. The trial arose from a collaboration between the Health Research Institute of Santiago de Compostela (IDIS), the Galician Health Service (SERGAS, through the General Directorate of Public Health), and the Galician Regional Vaccination Program.

Galicia’s healthcare system encompasses seven sanitary areas and 14 public hospital complexes, with a total of 9,818 beds (2022 data).^15^ All residents are assigned a unique SERGAS identification number, enabling linkage across care settings and facilitating nearly complete follow-up (>98% coverage).^16^ Population mobility in Galicia is low, reducing potential losses to follow-up.

In 2022, 26.3% of the Galician population was 65 years or older, and the number aged ≥85 years was 5.2%.^17^ During the 2022/2023 influenza season, HD-IIV became available for those aged ≥85 years and nursing home residents aged ≥60, while SD-IIV was standard for individuals aged 65–84.^11^ Vaccine uptake among those aged ≥65 exceeded 74%, nearing the WHO’s 75% coverage target, with especially high rates among those ≥80 years (86%) and slightly lower rates in the 65–69 age group (62%).^9,18^

### Study Design

GALFLU is a pragmatic, registry-based, open-label, active-controlled, individually randomized trial conducted over two consecutive influenza seasons (2023/24 and 2024/25). This design is especially well-suited to evaluating vaccines in older populations with varied comorbidity profiles. Baseline clinical characteristics, medication use, and prior vaccination status are defined in Supplementary Tables 1–3.

Each season, participants were randomized 1:1 to receive QIV-HD (Efluelda®) or standard quadrivalent SD-IIV (Influvac tetra®). Randomization was implemented centrally using a computer-generated block algorithm and embedded within routine vaccination operations. Individuals recruited in the first season could re-enroll with new randomization in the subsequent season. Enrollment was conducted from 26 October to 26 November in 2023/24, and from 30 October to 2 December in 2024/25, with follow-up for all endpoints beginning 14 days post-vaccination and continuing through May 31.

### Recruitment Strategy

Administrative registries and electronic health records facilitated identification of eligible participants, streamlined recruitment, and enabled comprehensive data collection throughout the study. Personalized SMS invitations with QR codes were used to streamline scheduling and participant identification at the point of vaccination. A custom software application supported efficient randomization by research staff. Each year, approximately 450,000 Galicians aged 65–79 years received initial invitations via SMS. To optimize participation, the study was supported by tailored outreach such as posters, informational leaflets, a web portal (www.galfu.gal), media releases, and a dedicated call center.

All recruitment, consent, and vaccination activities occurred within Galicia’s standard vaccination infrastructure. Enrollment occurred at 14 vaccination sites staffed by 190 sub-investigators coordinated by three regional leads. Potential participants could access full trial information before their appointment via SMS, web materials, in-person information, and a 24/7 staffed call center. Detailed trial information and downloadable informed consent forms were also provided at www.galfu.gal, with a frequently asked questions section and free hotline for additional inquiries.^19^

At the vaccination visit, patients expressing interest received a written information sheet and oral explanation by a trained medical professional, with opportunities to ask questions. Consent was obtained only after ensuring participant understanding and voluntary agreement. Those declining participation or withdrawing before vaccination could receive routine vaccination at the same visit and site.

### Ethics and Regulatory Oversight

The GALFLU protocol was approved by the Comité de ética de la investigación con medicamentos de Galicia (CEIm-G, code 2023/374) and registered under the EU Clinical Trials Regulation (536/2014) with oversight by the Spanish Agency of Medicines and Medical Devices (AEMPS). The study was conducted in accordance with the Declaration of Helsinki, ICH-GCP guidelines, and all applicable Spanish and European data protection laws (including GDPR and Organic Law 3/2018). Participation was fully voluntary, with the right to withdraw at any time. Data confidentiality was ensured: only pseudonymized data were transferred and handled under strict regulatory procedures.

### Inclusion Criteria

Owing to the pragmatic design, the only inclusion criteria were: (1) age 65–79 years; (2) residence in Galicia; (3) not eligible for HD-IIV as part of routine care; and (4) provision of informed consent. There were no additional exclusion criteria; vaccination contraindications were evaluated by the enrolling clinician as per local routine.

### Vaccine Administration

Vaccinations were delivered within the existing Regional Vaccination Program, according to product labeling and marketing authorizations. Co-administration of COVID-19 vaccines or boosters followed local guidance. Participants randomized to HD-IIV received Efluelda® (Sanofi, Paris); those randomized to SD-IIV received Influvac tetra® (Abbott Biologicals, The Netherlands). No blinding was implemented, given the nature of the intervention, but endpoints were ascertained objectively from health registries, minimizing risk of bias.

### Study Objectives and Endpoints

#### Primary Objective

To estimate the relative vaccine effectiveness (rVE) of HD-IIV versus SD-IIV against a composite endpoint of influenza- or pneumonia-related hospitalization in adults aged 65–79 years.

#### Secondary Objectives

To estimate rVE for HD-IIV versus SD-IIV against i) hospitalization for any cardio-respiratory disease as a composite endpoint; ii) all-cause hospitalization; iii) all-cause mortality; iv) hospitalization for influenza; and v) hospitalization for pneumonia.

#### Exploratory Objectives

Exploratory objectives included the rVE of HD-IIV versus SD-IIV against: (1) hospitalization for influenza or pneumonia as a composite endpoint (alternate definition); (2) hospitalization for pneumonia (alternate definition); (3) hospitalization for influenza (alternate definition); (4) hospitalization for any respiratory disease; (5) hospitalization for any cardiovascular disease; (6) hospitalization for myocardial infarction; (7) hospitalization for heart failure; (8) hospitalization for atrial fibrillation; (9) hospitalization for stroke; (10) major adverse cardiovascular events (MACE) defined as a composite of hospitalization for acute myocardial infarction, hospitalization for stroke, and cardiovascular death; (11) MACE defined as a composite of hospitalization for acute myocardial infarction, hospitalization for stroke, hospitalization for heart failure, and cardiovascular death (alternate MACE definition #1); (12) MACE defined as a composite of hospitalization for acute myocardial infarction, hospitalization for stroke, and all-cause death (alternate MACE definition #2); (13) hospitalization requiring mechanical ventilation; (14) laboratory-confirmed influenza; (15) hospitalization with a laboratory confirmed influenza; (16) laboratory-confirmed pneumococcal pneumonia; (17) laboratory-confirmed COVID-19; (18) cardio-respiratory mortality; (19) respiratory mortality; (20) cardiovascular mortality; and (21) in-hospital mortality for any cause.

#### Healthcare Resource Utilization Objectives

GALFLU aimed to assess healthcare resource use and related costs between HD-IIV and SD-IIV arms. Outcomes include: (1) any hospitalization; (2) hospitalizations requiring mechanical ventilation; (3) duration of hospitalization stays for: i) Influenza or pneumonia; ii) any cardio-respiratory disease; iii) Influenza and iv) pneumonia; (4) primary care visits; (5) nursing home admission following any all-cause hospitalization; and (6) anti-infective use.

### Endpoint Definition and Ascertainment

All endpoints are summarized in Table 1. Hospitalizations associated with a COVID-19 ICD-10 discharge code (B342, B972—any position) were excluded from endpoint definitions.

**Table 1.** Endpoints under primary, secondary, and exploratory objectives of the GALFLU study.

| Endpoint | ICD-10 codes | Diagnosis type | Inpatient, outpatient, or minimum required length of hospitalization and other criteria | Timeframe |
| --- | --- | --- | --- | --- |

| Primary objective endpoint |  |  |  |  |
| --- | --- | --- | --- | --- |
| Influenza or pneumonia hospitalization [composite endpoint] | J09-J18 | Primary discharge diagnosis | Inpatient, at least 1 night | ≥14 days after vaccination and up to May 31 the following year |
| Secondary objective endpoint |  |  |  |  |
| Cardio-respiratory disease hospitalization* [composite endpoint] | I11, I13, I20-I25, I30-I31, I33, I38-I42, I46-I50, I60-I69, I74, J00-J06, J09-J18, J20-J22, J40-J47, J80-J81, J85-J86, J96 | Primary discharge diagnosis | Inpatient, at least 1 night | ≥14 days after vaccination and up to May 31 the following year |
| All-cause hospitalization | Any | Any | Inpatient, at least 1 night | ≥14 days after vaccination and up to May 31 the following year |
| All-cause mortality | - | - | - | ≥14 days after vaccination and up to May 31 the following year |
| Influenza hospitalization | J09-J11 | Primary discharge diagnosis | Inpatient, at least 1 night | ≥14 days after vaccination and up to May 31 the following year |
| Pneumonia hospitalization | J12-J18 | Primary discharge diagnosis | Inpatient, at least 1 night | ≥14 days after vaccination and up to May 31 the following year |
| Exploratory objectives endpoints |  |  |  |  |
| Influenza or pneumonia hospitalization [composite endpoint] (alternate definition) | J09-J18 | Primary or secondary discharge diagnosis | Inpatient, at least 1 night | ≥14 days after vaccination and up to May 31 the following year |
| Laboratory-confirmed influenza hospitalization | - | Positive influenza microbiology test | Inpatient, at least 1 night | ≥14 days after vaccination and up to May 31 the following year |
| Influenza hospitalization (alternate definition) | J09-J11 | Primary or secondary discharge diagnosis | Inpatient, at least 1 night | ≥14 days after vaccination and up to May 31 the following year |
| Pneumonia hospitalization (alternate definition) | J12-J18 | Primary or secondary discharge diagnosis | Inpatient, at least 1 night | ≥14 days after vaccination and up to May 31 the following year |
| Respiratory disease hospitalization | J00-J06, J09-J18, J20-J22, J40-J47, J80-J81, J85-J86, J96 | Primary discharge diagnosis | Inpatient, at least 1 night | ≥14 days after vaccination and up to May 31 the following year |
| Cardiovascular disease hospitalization | I11, I13, I20-I25, I30-I31, I33, I38-I42, I46-I50, I60-I69, I74 | Primary discharge diagnosis | Inpatient, at least 1 night | ≥14 days after vaccination and up to May 31 the following year |
| Myocardial infarction hospitalization | I21 | Primary discharge diagnosis | Inpatient, at least 1 night | ≥14 days after vaccination and up to May 31 the following year |
| Heart failure hospitalization | I50 | Primary discharge diagnosis | Inpatient, at least 1 night | ≥14 days after vaccination and up to May 31 the following year |
| Atrial fibrillation hospitalization | I48 | Primary discharge diagnosis | Inpatient, at least 1 night | ≥14 days after vaccination and up to May 31 the following year |
| Stroke hospitalization | I63-I64 | Primary discharge diagnosis | Inpatient, at least 1 night | ≥14 days after vaccination and up to May 31 the following year |
| Major adverse cardiovascular events (MACE) | Hospitalizations: I21, I63-I64<br>Deaths: Any I-diagnosis as cause of death | For hospitalizations: Primary discharge diagnosis | For hospitalizations: Inpatient, at least 1 night | ≥14 days after vaccination and up to May 31 the following year |
| MACE (alternate definition #1) | Hospitalizations: I21, I50, I63-I64<br>Deaths: Any I-diagnosis as cause of death | For hospitalizations: Primary discharge diagnosis | For hospitalizations: Inpatient, at least 1 night | ≥14 days after vaccination and up to May 31 the following year |
| MACE (alternate definition #2) | Hospitalizations: I21, I63-I64<br>Deaths: All-cause | For hospitalizations: Primary discharge diagnosis | For hospitalizations: Inpatient, at least 1 night | ≥14 days after vaccination and up to May 31 the following year |
| Hospitalization requiring mechanical ventilation | - | - | Any duration, procedural code "BGDA" registered at any point during hospitalization | ≥14 days after vaccination and up to May 31 the following year |
| Laboratory-confirmed pneumococcal pneumonia | - | - | Positive test for streptococcus pneumonia | ≥14 days after vaccination and up to May 31 the following year |
| Laboratory-confirmed COVID-19 | - | - | Positive COVID-19 test | ≥14 days after vaccination and up to May 31 the following year |
| In-hospital mortality | - | - | Death (all-cause) while being hospitalized for at least 1 night | ≥14 days after vaccination and up to May 31 the following year |
| Cardio-respiratory mortality | Any I- or J-diagnosis as a cause of death | - | - | ≥14 days after vaccination and up to May 31 the following year |
| Respiratory mortality | Any J-diagnosis as a cause of death | - | - | ≥14 days after vaccination and up to May 31 the following year |
| Cardiovascular mortality | Any I-diagnosis as cause of death | - | - | ≥14 days after vaccination and up to May 31 the following year |

*\*: For cardio-respiratory hospitalizations, a broad list of ICD-10 codes is currently specified. ICD: International Classification of Diseases.*

### Sample Size Calculation

A minimum sample size of 114,011 participants (approximately 57,000 per season) was planned to provide ≥80% power to detect an rVE of at least 18% (the local cost-effectiveness threshold), assuming a one-sided Type I error of 0.025, a 1:1 allocation ratio, and a 0.8% event rate for the primary endpoint in the SD-IIV group (based on latest Galician registry data).

### Data Sources and Linkage

All data—including baseline characteristics, exposures, endpoints, and healthcare resource utilization—were obtained from the SERGAS registries and associated administrative databases (see Supplementary Text 1). Each participant was assigned a unique study code at enrollment and randomization, and identifiers were securely stored on a restricted-access list.

### Safety Monitoring

Both trial vaccines were approved and monitored for safety using a registry-based approach. An independent committee evaluated occurrence of serious adverse events (SAEs) and serious adverse reactions (SARs) through three months post-vaccination (±15 days), using automated extraction from health registries (see Supplementary Text 2).

### Statistical Analysis Plan

Continuous baseline variables will be summarized with means and standard deviations or medians and interquartile ranges; categorical variables will be summarized as frequencies and proportions. p-values <0.05 will indicate statistical significance, with alpha-adjusted thresholds as required for multiple comparisons.

For all rVE outcomes, only the participant’s first qualifying event per endpoint category will be considered: rVE = (1 - (CHD/NHD) / (CSD/NSD)) × 100%, where CHD and CSD denote the number of endpoints in HD-IIV and SD-IIV arms, and NHD, NSD are the numbers randomized to each arm. Confidence intervals (95% alpha-adjusted) will use the Clopper–Pearson method. p-values will only be reported for endpoints tested in the hierarchical sequence; inference will be based on the lower CI. Superiority of HD-IIV over SD-IIV will be concluded if the lower rVE CI is >0%.

Planned sensitivity analyses include varying the follow-up period (e.g., extending through late summer or restricting to peak influenza activity), incorporating time-to-event analyses with Cox regression and log-rank tests, examining within-person correlation across seasons, and alternate approaches to endpoint definitions (see Supplementary Text 3).

### Operational Implementation

#### Staff Training and Preparation

During the first GALFLU season, 181 research nurses underwent comprehensive training on all study procedures. This included a general site initiation visit and 14 on-site training sessions conducted at vaccination clinics. All study team members completed Good Clinical Practice (GCP) training to ensure compliance with regulatory and ethical standards. Training covered participant recruitment, consent, vaccination procedures, data collection, and randomization.

For the second season, additional nurses were recruited and trained, expanding the workforce to 187 trained research nurses. Many of these nurses benefited from the expertise of first-season staff, enhancing continuity and operational knowledge.

### Recruitment Planning and Target Setting

Recruitment objectives were set based on historical influenza vaccination coverage within Galicia and an estimated trial participation rate of approximately 17.5%. The overall enrollment target was approximately 57,000 participants per season, distributed across the 14 vaccination sites. Site-specific recruitment goals were derived to reflect regional vaccination rates and population distribution.

These targets were closely monitored throughout the recruitment period to ensure balanced enrollment and optimal geographic coverage.

### Digital Infrastructure and Randomization Tools

A bespoke randomization application was developed to support the trial’s operational needs. This application allowed multiple users at different vaccination sites to perform rapid, simultaneous individual randomizations, seamlessly integrated within the routine vaccination workflow. It facilitated participant registration, vaccine allocation, and real-time data capture without interrupting clinical flow.

The digital platform proved essential in achieving high throughput and minimizing delays during vaccination appointments.

### Participant Invitation and Engagement

Approximately 450,000 eligible Galicians aged 65–79 years were invited to participate each season via personalized SMS containing QR codes. These QR codes linked participants directly to the recruitment and scheduling system, simplifying the vaccination appointment process.

To raise awareness and encourage participation, additional outreach included:

- Informational posters and leaflets prominently displayed at vaccination centers and public areas
- An informative website (www.galfu.gal) featuring trial details, downloadable consent forms, and a FAQ section
- Media releases and local news coverage
- A 24/7 dedicated call center staffed by trained personnel responding to participant inquiries, clarifying trial processes, and assisting with appointment scheduling

### Site Coordination and Support

Each vaccination site was staffed with research nurses who were supported by nurse coordinators responsible for overseeing daily recruitment operations and maintaining protocol fidelity. These site coordinators liaised with the central GALFLU coordination team, which provided overarching guidance, operational assistance, and troubleshooting support.

The central coordination team was available around the clock during recruitment periods, ensuring rapid responses to any issues encountered at sites.

### Recruitment Timelines and Performance Metrics

Recruitment for the first season (2023/24) began on 26 October 2023 and the overall target of 57,326 participants was reached within 24 days. Recruitment continued for an additional 6 days, culminating in the enrollment of 59,490 participants.

Twelve of the fourteen vaccination sites met their recruitment goals within the planned period. The fastest site reached its target in 16 days. Peak recruitment reached 4,136 participants per day.

During the second season (2024/25), recruitment performance improved following operational refinements. The recruitment goal of 57,036 participants was met even faster—by day 19—and recruitment continued beyond the target to enroll a total of 74,986 participants (131.5% of the goal). The fastest site during the second season achieved the target in just 12 days.

Daily recruitment rates were consistently higher throughout the second season compared to the first, demonstrating increased efficiency and scale.

### Operational Challenges and Solutions

Throughout recruitment, no major issues or interruptions in the randomization or vaccination processes were reported. The study team’s readiness, combined with the digital randomization tool and comprehensive staff training, minimized operational risks.

Where minor logistical bottlenecks arose, such as fluctuations in participant flow or appointment scheduling, the centralized coordination team quickly intervened to adjust resources and workflows.

### Improvements and Adaptations for the Second Season

Lessons learned from the first season informed logistical and human resource improvements before the second season. Enhancements included:

- Expanded and targeted research nurse training programs, ensuring additional staffing capacity
- Streamlined appointment scheduling and participant processing
- Refinements to the digital randomization platform to handle increased enrollment volumes efficiently
- Strengthened communication efforts to optimize participant engagement and satisfaction

These adaptations directly contributed to improved recruitment speed, higher enrollment rates, and the achievement of recruitment goals at all sites during the second season.

## Discussion

GALFLU, to the best of our knowledge, is the first adequately powered pragmatic, individually randomized trial to assess the comparative effectiveness of HD-IIV against SD-IIV for a comprehensive range of clinically important endpoints, including severe hospitalizations, in a general population of adults aged 65–79 years.

Annual influenza vaccination remains the most effective means of reducing disease burden in the elderly. However, lower immunogenicity and clinical effectiveness in this age group have driven the development and preferential recommendation of HD-IIV formulations. While evidence from controlled trials supports improved immunogenicity, policy determinations require robust real-world effectiveness data.

The pragmatic design of GALFLU—encompassing broad eligibility, integration within the existing health infrastructure, streamlined digital recruitment and follow-up, and objective, registry-based endpoint ascertainment—ensures high external validity. Additional personnel were hired for operational efficiency, reflecting potential real-world resource investments required for rapid trial deployment. Importantly, the trial preserved the minimal-disruption ethos of pragmatic methodology: no extra screening visits, research-specific contacts, or data collection burdens on participants.

Innovations in digital recruitment (personalized SMS, QR codes), public health messaging, and the use of routine registry data have demonstrated a scalable, low-cost model for integrating research into practice. Such approaches enhance feasibility and acceptability and may accelerate progress toward learning health systems—where health care delivery and evidence generation are dynamically integrated.

GALFLU’s operational success demonstrates that pragmatic, registry-based, individually randomized trials can be conducted efficiently and at scale, with high data completeness and minimal attrition. This sets a precedent not only for influenza vaccine evaluations, but for comparative effectiveness research embedded within any established public health program.

## Conclusions

GALFLU demonstrates the high feasibility and scalability of pragmatic, individually randomized influenza vaccine trials within routine care for older adults in Galicia. This operational framework, leveraging digital infrastructure and population registries, enables robust, population-level evidence on vaccine effectiveness and has direct implications for regulatory and policy decision-making. The trial provides a reproducible blueprint for future pragmatic evaluations embedded in routine public health settings. Final study results will be reported separately.

## Supporting information

annex GALFLU team

## Data Availability

Results produced will be published.

## Trial Registration

EU Clinical Trials Register (2023-506977-36-00); ClinicalTrials.gov (NCT06141655)

## Key points

GALFLU shows the feasibility of large, pragmatic, individually randomized trials and highlights the value of population-based health registries embedded within public health systems, offering randomized real-world evidence to inform vaccine policy, regulatory decisions, and cost-effectiveness analyses for influenza and other vaccines.

