## Supplementary Text 1 for "Methods and Operational Framework of the GALFLU Pragmatic Trial: Design and Feasibility of an Individually Randomized Controlled Trial Evaluating High-Dose Versus Standard-Dose Influenza Vaccination in Older Adults"

**Data sources**

The data were collected using the following SERGAS registries: i) Minimum Basic Data Set (CMBD, Conjunto Mínimo Básico de Datos) which includes hospitalization data – e.g. ICD 10 codes (discharge diagnosis), in-hospital mortality, and length of hospitalization, among others; ii) health card registry which includes personal identification data such as age, sex and residence address; iii) Galician registry of mortality provides data on date and cause of death; iv) Galician Information Hospitalization System (SIHGA, Sistema información hospitalaria de Galicia) which encompass data on all-cause hospitalization, hospitalization contacts (such as date of admission, date of discharge and in-hospital mortality); v) clinical cards registry on morbidity associated with the most prevalent chronic diseases; vi) primary care visits registry; vii) vaccines registries (RVACU); viii) medicines discharge registry (ATC codes); ix) microbiology registry (laboratory-confirmed respiratory infections such as influenza, COVID-19, and pneumococcus, among others). Additional information about potential safety events experienced by trial participants is collected from medical records.
