## Supplementary Text 2 for "Methods and Operational Framework of the GALFLU Pragmatic Trial: Design and Feasibility of an Individually Randomized Controlled Trial Evaluating High-Dose Versus Standard-Dose Influenza Vaccination in Older Adults"

**Safety assessment**

Both the QIV-HD and QIV-SD used in the GALFLU trial were approved for use in Galicia. Accordingly, safety was monitored using a pragmatic, registry-based approach. Each participant was assessed for the occurrence of serious adverse events (SAEs) and serious adverse reactions (SARs) at three months after vaccination (+/- 15 days) using data extracted from SERGAS registries.

A list of study participants who experienced a SAE was generated using a registry-based program designed for this purpose. The execution of the program, processing of the raw registry data, and production of the list for the safety assessment were carried out by the research team in a closed remote-access environment hosted on an encrypted server of the Galician Health Data Authority, which contained the raw registry data. The causality of each SAE with the study treatment was determined by a medical doctor from the research team based on a review of the electronic medical records.

If an SAE occurred under the following conditions: (i) the participant was hospitalized at the time of SAE assessment or the SAE had not yet been resolved, and (ii) a medical doctor determined that the hospitalization was causally unrelated to the study treatment, then the event was recorded as an SAE, and no further follow-up on that specific incident was required. Similarly, if (i) a participant was hospitalized at the time of SAE assessment or the SAE had not yet been resolved; (ii) a medical doctor determined that the SAE incident was causally related to the study treatment, and (iii) the incident was a side effect already mentioned in the product summary, then the event was recorded as a SAR, and no further follow-up on that particular incident was conducted.

In both cases, only the participant's identification number and the date of the SAE or SAR were recorded in the electronic case report form.

The product summaries of the QIV-HD vaccine and the comparator QIV-SD vaccine(s) were used to determine whether a SAR is unexpected and thus possibly a suspected unexpected SAR (SUSAR). Life-threatening and fatal SUSARs were reported to the Spanish Agency for Drugs and Health at the earliest time point and no later than seven days after the incident registration. All other SUSARs were reported to the Ethics Committee within 15 days. The number of SAEs, SARs, and SUSARs was reported, in both annual and final reports, to the Spanish Agency for Drugs and Health and the Ethics Committee.
