## Supplementary Text 3 for "Methods and Operational Framework of the GALFLU Pragmatic Trial: Design and Feasibility of an Individually Randomized Controlled Trial Evaluating High-Dose Versus Standard-Dose Influenza Vaccination in Older Adults"

**Sensitivity analysis**

Several sensitivity analyses will be performed to assess the robustness of the rVE estimates.

***Extended follow-up period***

An extended follow-up period will be considered, covering May 31 to August 31 of each influenza season. This extended window is designed to capture additional downstream events potentially attributable to influenza activity occurring later in the season.

***Restricted follow-up period***

To focus the analysis on periods of heightened influenza activity, two distinct approaches will be used to define a restricted follow-up period.

Surveillance-based approach: Using national Galician influenza surveillance data, periods of viral activity will be defined as follows: i) Moderate/high activity: From the first to the last week (inclusive) during which influenza test positivity exceeds the 55th percentile; and ii) High Activity: From the first to the last week (inclusive) during which influenza test positivity exceeds the 75th percentile.

Hospitalization-based approach: the influenza peak period is defined according to influenza or pneumonia hospital admission rates. The reference point is the week with the highest incidence of influenza or pneumonia hospitalizations among individuals aged 65 to 79 years in Galicia. The observation period will extend from two weeks before to six weeks after this peak week. The incidence rate will be assessed using registry data and the same prespecified endpoint definitions.

***Survival analysis***

In this sensitivity analysis, all effectiveness endpoints will be analyzed using survival analysis instead of calculation of rVE. Only the participants’ first event in each endpoint category will be considered. Crude Cox proportional hazards regression will be used to calculate hazard ratio (HR) and 95% CI for each endpoint. p-values will be generated using a log-rank test.

***No exclusion of endpoints with associated COVID-19 diagnosis codes***

In this sensitivity analysis, endpoints with associated COVID-19 discharge diagnosis codes will not be excluded. This sensitivity analysis will only be conducted for the primary and secondary endpoints.

***Analysis accounting for potential within-participant correlation across seasons***

An analysis of the primary and secondary endpoints based on a model accounting for potential within-participant correlation in participants enrolled in >1 season.

***Follow-up period starting from time of randomization***

In this sensitivity analysis, the follow-up period will include the 14-day interval from time of vaccination that is excluded in the endpoint definitions meaning that the follow-up period for this sensitivity analysis will be defined as from the time of randomization until May 31 the following year.
