## Supplementary Table 1 for "Methods and Operational Framework of the GALFLU Pragmatic Trial: Design and Feasibility of an Individually Randomized Controlled Trial Evaluating High-Dose Versus Standard-Dose Influenza Vaccination in Older Adults"

**Supplementary Table 1. Prespecified definitions for baseline medical conditions of the GALFLU study participants**

| **Condition** | **ICD-10/ATC codes*** | **Diagnosis type** | **Inpatient, outpatient, minimum required length of hospitalization, other criteria** | **Timeframe (before randomization)** |
| --- | --- | --- | --- | --- |
| No. of hospitalizations in the last 12 months | Any | Any | Inpatient, at least 1 night | ≤12 months |
| No. of hospitalizations in the last 10 years | Any | Any | Inpatient, at least 1 night | ≤10 years |
| No. of hospital contacts in the last 12 months | Any | Any | Inpatient, at least 1 night | ≤12 months |
| No. of hospital contacts in last 10 years | Any | Any | Inpatient, at least 1 night | ≤10 years |
| No. of primary care contacts in the last 12 months | Any | Any | - | ≤10 years |
| No. of primary care contacts in the last 10 years | Any | Any | - | ≤10 years |
| Chronic obstructive pulmonary disease | J42-J44 | Primary or secondary discharge diagnosis | Any | ≤10 years |
| Asthma | J45 | Primary or secondary discharge diagnosis | Any | ≤10 years |
| Chronic lung disease | A15-A16, D860, E84, J42-J47, J84, Z942 | Primary or secondary discharge diagnosis | Any | ≤10 years |
| Diabetes (ICD-10) | E10-E14 | Primary or secondary discharge diagnosis | Any | ≤10 years |
| Diabetes (ATC) | A10 | - | ≥1 claimed prescription | ≤180 days |
| Hypertension (ICD-10) | I10-I15 | Primary or secondary discharge diagnosis | Any | ≤10 years |
| Hypertension (ATC) | Diuretics: C03  -blockers: C07  Calcium blockers: C08  Renin-angiotensin system inhibitors: C09 | - | ≥1 claimed prescription from ≥2 drug classes | ≤180 days |
| Dyslipidemia (ATC) | C10 | - | ≥1 claimed prescription | ≤180 days |
| Ischemic heart disease | I20-I25 | Primary or secondary discharge diagnosis | Any | ≤10 years |
| Myocardial infarction | I21 | Primary or secondary discharge diagnosis | Any | ≤10 years |
| Heart failure | I50 | Primary or secondary discharge diagnosis | Any | ≤10 years |
| Other cardiomyopathy | I42-I43 | Primary or secondary discharge diagnosis | Any | ≤10 years |
| Atrial fibrillation | I48 | Primary or secondary discharge diagnosis | Any | ≤10 years |
| Other arrhythmia | I44-I47, I49 | Primary or secondary discharge diagnosis | Any | ≤10 years |
| Valvular disease | I34-I37 | Primary or secondary discharge diagnosis | Any | ≤10 years |
| Pericarditis | I30-I31 | Primary or secondary discharge diagnosis | Any | ≤10 years |
| Endocarditis | I33, I38-I39 | Primary or secondary discharge diagnosis | Any | ≤10 years |
| Myocarditis | I40-I41 | Primary or secondary discharge diagnosis | Any | ≤10 years |
| Pulmonary heart disease | I26-I28 | Primary or secondary discharge diagnosis | Any | ≤10 years |
| Cerebrovascular disease | I60-I69 | Primary or secondary discharge diagnosis | Any | ≤10 years |
| Peripheral vascular disease | I70, I74 | Primary or secondary discharge diagnosis | Any | ≤10 years |
| Congenital heart disease | Q20-Q26 | Primary or secondary discharge diagnosis | Any | ≤10 years |
| Chronic cardiovascular disease | I20-I28, I34-I37, I42-I50, I60-I69, I70, I74, Q20-Q26 | Primary or secondary discharge diagnosis | Any | ≤10 years |
| Cancer | C00-C97 (not C44) | Primary or secondary discharge diagnosis | Any | ≤10 years |
| Chronic kidney disease | E102, E112, E132, E142, I120, N02-N08, N11-N12, N14, N18-N19, N26, N158-N160, N162-N164, N168, M300, M313, M319, M321B, Q612-Q613, Q615, Q619, T858-T859, Z992 | Primary or secondary discharge diagnosis | Any | ≤10 years |
| Liver disease | B15-B19, K70-K77, C22, I982, Z944, D684C, Q618 | Primary or secondary discharge diagnosis | Any | ≤10 years |
| Immunodeficiency (ICD-10) | B20-B24, D80-D84, D89, O987, Z21, Z940-Z944, Z948A | Primary or secondary discharge diagnosis | Any | ≤10 years |
| Immunodeficiency (ATC) | H02AB, L04 | - | ≥1 claimed prescription | ≤180 days |
| Neurological/  neuromuscular disease | F00-F03, G10-G14, G20-G23, G30-G32, G35-G37, G40-G41, G70-G73, G80-G83, G91-G95 | Primary or secondary discharge diagnosis | Any | ≤10 years |
| Dementia | F00-F03, G30, G311-G312, | Primary or secondary discharge diagnosis | Any | ≤10 years |
| Rheumatic disease | M05-M06, M32-M34, M353 | Primary or secondary discharge diagnosis | Any | ≤10 years |
| Anemia | D50, D62, D64 | Primary or secondary discharge diagnosis | Any | ≤10 years |
| Presence of at least one chronic disease | At least one of the following:  Chronic lung disease; Diabetes; Chronic cardiovascular disease;  Cancer;  Chronic kidney disease;  Immunodeficiency;  Neurological/neuromuscular disease;  Liver disease;  Rheumatic disease | - | - | - |

**: All baseline conditions are defined from ICD-10 codes except for diabetes, hypertension, dyslipidemia, and immunodeficiency, where ATC codes are also used. ICD: International Classification of Diseases; ATC: Anatomic Therapeutic Chemical.*
