## Supplementary Table 2 for "Methods and Operational Framework of the GALFLU Pragmatic Trial: Design and Feasibility of an Individually Randomized Controlled Trial Evaluating High-Dose Versus Standard-Dose Influenza Vaccination in Older Adults"

**Supplementary Table 2. Prespecified definitions for medication use (at least one claimed prescription) at baseline for the GALFLU participants. Medication use at baseline will be assessed using the Anatomic Therapeutical Chemical (ATC) codes. A timeframe ≤180 days before randomization was established.**

| **Medication** | **ATC codes** |
| --- | --- |
| Antithrombotics | B01 |
| Renin-angiotensin system inhibitor | C09 |
| Calcium blockers | C08 |
| Beta-blockers | C07 |
| Diuretics | C03 |
| Aspirin | B01AC06 |
| Statins | C10AA |
| Inhaled beta-2 agonists | R03A |
| Inhaled anticholinergics | R03BB, R03AL01-07 |
| Inhaled glucocorticoids | R03BA, R03AK, R03AL08-09, R03AL11-12 |
| Insulin | A10A |
| Non-insulin antidiabetic medication | A10B |
| Systemic glucocorticoids | H02AB |
| Immunosuppressants | L04 |
