## Supplementary Table 3 for "Methods and Operational Framework of the GALFLU Pragmatic Trial: Design and Feasibility of an Individually Randomized Controlled Trial Evaluating High-Dose Versus Standard-Dose Influenza Vaccination in Older Adults"

**Supplementary Table 3. Prespecified definitions for previous vaccinations at baseline for the GALFLU participants. Previous vaccinations will be determined using the Anatomic Therapeutical Chemical (ATC) codes. Each vaccination will have specific assessment criteria.**

| **Medication** | **ATC codes** | **Assessed as** |
| --- | --- | --- |
| Influenza vaccination in the previous season | J07BB | Vaccinated in the previous season (October 1 through May 31) |
| No. of influenza vaccinations in the past 5 seasons | J07BB | No. of vaccinations in the past 5 seasons (assessed between October 1 through May 31 for each season) |
| Previous pneumococcal vaccination | J07AL | At least 1 vaccine after the age of 65 |
| Previous COVID-19 vaccination | J07BX03 | At least 1 vaccine received at any time before inclusion |
