## Supplementary material for "Methods and Operational Framework of the GALFLU Pragmatic Trial: Design and Feasibility of an Individually Randomized Controlled Trial Evaluating High-Dose Versus Standard-Dose Influenza Vaccination in Older Adults": annex GALFLU team

**GALFLU trial investigator team**

**The GALFLU study team was composed by, in alphabetical order:**

| **Name and surname** | **Affiliation** |
| --- | --- |
| Ana Maria Abal Senin | 1 |
| Pilar Alonso Vence | 1 |
| Rosa María Álvarez Gil | 1 |
| Talia Alvarez Suarez | 1 |
| Lidia Alvarez Dapia | 1 |
| Jose Manuel Alvarez Pena | 1 |
| Silvia Amoedo Casqueiro | 1 |
| Patricia Aran Abelenda | 1 |
| Josefa Araujo Rodriguez | 1 |
| Ana Ares Martinez | 1 |
| Maria Asensio Nieto | 1 |
| Noelia Belen Ayllon Morcillo | 1 |
| Mónica Barral Carregal | 1 |
| Maria Begoña Baquero Sabajanes | 1 |
| Alejandro Baña Tomé | 1 |
| Lorena Bayon Diz | 1 |
| Xavier Bello Paderne | 2 |
| Miriam Ben García | 2 |
| Damian Bernardez Sende | 1 |
| Beatriz Blanco Salgado | 1 |
| Lucia Bruquet Costa | 1 |
| Faustino Caballero Muinelo | 1 |
| Noelia Cabeiras Alvarez | 1 |
| Maria Guadalupe Cainzos Fernandez | 1 |
| Alba Camino Mera | 2 |
| Alba Campero Ojea | 1 |
| Isabel Campos Pardo | 1 |
| Diego Canedo Cotelo | 1 |
| Sandra Carnota Antonio | 2 |
| Lidia Carreira Diaz | 1 |
| Miguel Angel Carreiro Alonso | 1 |
| Almudena Carro Orgeira | 1 |
| Adriana Maria Castelo Triñanes | 1 |
| Ana Belen Castelo Vazquez | 1 |
| César Castro Pita | 1 |
| Andrea Castro Sande | 1 |
| Manuel Castro Guizan | 1 |
| Mercedes Castro Bacariza | 1 |
| Maria Jose Castro Castro | 1 |
| Maria Jose Castroman Souto | 1 |
| Sabela Ceide Fernandez | 1 |
| Silvia Cerqueiro Rey | 2 |
| Olga María Ces Ozores | 1 |
| Rebeca Chouza Alonso | 1 |
| Verónica Civeira Pérez | 1 |
| Carla Alexandra Correia Parauta | 1 |
| Ana Isabel Corbal Obelleiro | 1 |
| Ana Cotovad Bellas | 2 |
| Angela Coutado Belon | 1 |
| Xurxo Couto Catoira | 1 |
| Ana Isabel Couto Tobío | 1 |
| Evaristo Cuerdo Coton | 1 |
| Angeles Cuiña Caneda | 1 |
| María José Currás Tuala | 2 |
| Ana Dacosta Urbieta | 2 |
| Sonia De Miguel Cañas | 1 |
| Paola De Castro Garcia | 1 |
| Maria Eugenia De Jesús Torrealba Valente | 1 |
| Carmen Del Ojo Paez | 1 |
| María Begoña Del Oro Saez | 1 |
| Romina Deus Suarez | 1 |
| Paula Diaz Lopez | 1 |
| Lucia Diaz Gonzalez | 1 |
| Alejandro Docal Pombo | 1 |
| Marta Dominguez Lago | 1 |
| Beatriz Dominguez Picos | 1 |
| Rita Dominguez Gonzalez | 1 |
| Eva Maria Dominguez Martis | 1 |
| Maria Eulalia Dominguez Vilas | 1 |
| Ángeles Dono Díaz | 1 |
| Olalla Duro Liñares | 1 |
| Olalla Díaz Zuaza | 1 |
| Eloy Díez Polo | 1 |
| Estefania Esmoris Suarez | 1 |
| Rocio Estepa Diaz | 1 |
| Maria Montserrat Estevez Fernandez | 1 |
| Gemma Fandiño Guardado | 1 |
| Veronica Fernandez Gronewold | 1 |
| Noemi Fernandez Martinez | 1 |
| Alfonso Fernandez Vazquez | 1 |
| Susana Fernandez Perez | 1 |
| Raquel Fernandez Rodriguez | 1 |
| Nedea Fernandez Rama | 1 |
| Natalia Fernandez Novelle | 1 |
| Sebastian Estebo Fernández De Las Heras | 1 |
| Marta Fernandez Del Rio | 1 |
| Cristina Fernandez Costa | 1 |
| Anselmo Fernandez Alonso | 1 |
| Monica Beatriz Fernandez Martin | 1 |
| Maria Pilar Fernandez Varela | 1 |
| Isabel Ferreiros Vidal | 2 |
| Rosa Maria Ferro Castaño | 1 |
| Andrea Filgueira Vazquez | 1 |
| Melania Fraga Deus | 1 |
| Milagros Valeria Franco Soñora | 1 |
| Mercedes Fuentes Fernandez | 1 |
| Margarita Galdo Sierra | 1 |
| Verónica Garcia Romero | 1 |
| Carmen Garcia Garcia | 1 |
| Cecilia García Duran | 1 |
| Maria Gestoso Filloy | 1 |
| Maria De La Cruz Gil Rodriguez | 1 |
| Iago Giné Vázquez | 2 |
| Laura Giraldez Groba | 1 |
| Alberto Gómez Carballa | 2 |
| Dorotea Gomez Fernandez | 1 |
| Cristina Gomez Lendoiro | 1 |
| Cristina Gomez Lopez | 1 |
| Alejandro Gomez Moldes | 1 |
| Miguel Gonzalez Moron | 1 |
| Jose Gómez Rial | 2 |
| Laura Gonzalez Carballo | 1 |
| Natalia González Cacho | 1 |
| Maria Gonzalez Fondado | 1 |
| Ana González Isorna | 1 |
| Maria Mar Gonzalez Mendez | 1 |
| Maria Carmen Gonzalez Nisarre | 1 |
| Marina Gonzalez Zar | 1 |
| Cristian Guerreiro Seijas | 1 |
| Alberto Gómez Carballa | 1 |
| Jose Gómez Rial | 1 |
| M.ª del Carmen Guerra Calvelo | 1 |
| Maria Angeles Guzman Ruiz | 1 |
| Maria Luisa Hermida Sanchez | 1 |
| Aida Hermida Trujillo | 1 |
| Maria Belén Hernández González | 1 |
| Laura Iglesias Carballo | 1 |
| Nuria Iglesias Canedo | 1 |
| Rita Iglesias Estevez | 1 |
| Maria Insua Lago | 1 |
| Lorena Íñiguez Tejo | 1 |
| Sara Jacobo Vazquez | 1 |
| Antia Jarazo Miras | 1 |
| Ana Jurjo Costa | 1 |
| Rosa Mª Lage Fontarigo | 1 |
| María Isabel Liñeira Mourón | 1 |
| Maria Del Cielo Lista Rodriguez | 1 |
| Fernando Lois Vidal | 1 |
| Amaya Lois Lama | 1 |
| Carmen Lopez Bolaño | 1 |
| Luz Maria Lopez Castro | 1 |
| Rosa Maria Lopez Cordero | 1 |
| Montserrat López Franco | 2 |
| Maria Begoña Lopez Garcia | 1 |
| Rocio Lopez Lopez | 1 |
| Miguel Lopez Potes | 1 |
| Laura Lopez Rodriguez | 1 |
| Ana Lorenzo Olmo | 1 |
| Lucia Lorenzo Quintans | 1 |
| Luis Enrique Lorenzo Vila | 1 |
| Paula Losada Del Rio | 1 |
| Sabela Losada Dieguez | 1 |
| Syra Maceira Guisamonde | 1 |
| Narmeen Mallah | 2 |
| Andrea Mallon Gonzalez | 1 |
| Ángela Manzanares Casteleiro | 2 |
| Gemma M.ª Marquina Diaz | 1 |
| Ana Maria Millan Agrasar | 1 |
| Nuria Martinez Blanco | 1 |
| Ángela Martinez Martinez | 1 |
| Lara Martínez Martínez | 2 |
| Estrella Martínez Moreira | 1 |
| Olga Martínez Regueira | 1 |
| Federico Martinón-Torres | 1,2 |
| Karin Meier Cacharo | 1 |
| Diego Mella Tembrás | 1 |
| Raquel Mera Amoedo | 1 |
| Maria Pilar Mera Rodríguez | 1 |
| Fernando Mercador Mouce | 1 |
| Cristina Miguez Torres | 1 |
| Rosa Ana Montaña Rodriguez | 1 |
| Esther Montero Campos | 2 |
| Julian Montoto Louzao | 2 |
| Carla Maria Morales Soliño | 1 |
| Jimena Moreira Barros | 1 |
| Maria Concepcion Morigosa Galiano | 1 |
| Iria Mosquera Flores | 1 |
| Belén Mosquera Pérez | 2 |
| Mar Mosteiro Valiño | 1 |
| Maria Muiña Rico | 1 |
| Maider Muiños Blanco | 1 |
| Andres Muy Pérez | 2 |
| Victoria Nartallo Penas | 1 |
| Laura Navarro Ramón | 2 |
| Ursula Nieto Camaño | 1 |
| Teresa Noguerol Gonzalez | 1 |
| Sabela Noval Couceiro | 1 |
| Ana Noya Gonzalez | 1 |
| Maria Elisa Nuñez Losada | 1 |
| Juan Ocon Casal | 1 |
| Pilar Osende Barallobre | 1 |
| María Teresa Otero Barros | 1 |
| Paula Otero Riveira | 2 |
| Iria Otero Tenorio | 1 |
| Paula Otero Riveira | 1 |
| Irene Otero Lopez | 1 |
| Veronica Oubiña Cousido | 1 |
| Jesus Padilla Castillo | 1 |
| Rosa Maria Padin Gonzalez | 1 |
| Eva Padrón Lloves | 1 |
| Aurora Pan Regueira | 1 |
| Silvia Parada Gañete | 1 |
| Jacobo Pardo Seco | 2 |
| Maria José Parrado Alonso | 1 |
| Ana María Pastoriza Mourelle | 2 |
| Mercedes Pedreira Dominguez | 1 |
| Sandra Maria Pedreira Perez | 1 |
| Maria Del Carmen Pereiro Belay | 1 |
| Ana Perez Godas | 1 |
| Carmen Perez Lourido | 1 |
| Maria Perez Neira | 1 |
| Eva Maria Perez Pena | 1 |
| Lara Perez Rodriguez | 1 |
| Nerea Perez Salgado | 1 |
| Ines Perez Sabin | 1 |
| Marta Piedra Viqueira | 1 |
| Dora Pintos Rodriguez | 1 |
| Sara Pischedda | 2 |
| Carmen Portela Sanmartin | 1 |
| Maria Beatriz Prego Garcia | 1 |
| Adrian Prego Del Rio | 1 |
| Ángela Pérez Vazquez | 1 |
| Félix Pérez Herráiz | 1 |
| Olaia Pérez Martínez | 1 |
| Iva Vanusa Pires Reis | 1 |
| Diego Rama Cordeiro | 1 |
| Andrea Raposo Varela | 1 |
| Josefina Razzini | 2 |
| Lorenzo Redondo Collazo | 2 |
| Alejandro Redondo Gerpe | 1 |
| Sonia Regueiro Amoedo | 1 |
| Patricia Regueiro Casuso | 2 |
| Jose Luis Rey Fole | 1 |
| Fátima Rey Trigo | 1 |
| Susana Rey García | 2 |
| Thania Rey Villamea | 1 |
| Sara Rey Vázquez | 2 |
| Barbara Rial Seijas | 1 |
| Irene Rivero Calle | 2 |
| Pablo Robledo Casado | 1 |
| Gemma Rodriguez Alvarez | 1 |
| Carmen Rodriguez Calvo | 1 |
| Susana Rodriguez Carballo | 1 |
| Celia Rodriguez Estoquera | 1 |
| Diana Rodriguez Loureiro | 1 |
| Antonio Rodriguez Pena | 1 |
| Digna Rodriguez Romar | 1 |
| Iria Rodriguez Romar | 1 |
| Alberto Rodriguez Seoane | 1 |
| Cesar Rodriguez Serrada | 1 |
| Susana Rodriguez Simon | 1 |
| Carmen Rodríguez-Tenreiro Sánchez | 2 |
| Ana Isabel Romero Pintos | 1 |
| Olaya Rosende Pernas | 1 |
| Alejandra Roura Gomez | 1 |
| Adrian Rubio Montalban | 1 |
| Alberto Salmonte Rodriguez | 1 |
| Alba Sanchez Casas | 1 |
| Maria Isolina Santiago Perez | 1 |
| Maria Begoña Santos Padin | 1 |
| Jessica Seco Franco | 1 |
| Ana M.ª Senín Ferreiro | 2 |
| Victoria Senlle Costas | 1 |
| Sonia Serén Fernández | 2 |
| Melina Soengas Gonzalez | 1 |
| Susana Soliño Lourido | 1 |
| Laura Somoza Copa | 1 |
| Begoña Soriano Lorente | 1 |
| Maria José Soriano Ureña | 1 |
| Norma Suarez Castro | 1 |
| Nuria Suárez Gaiche | 1 |
| Pablo Taboada Pampin | 1 |
| Miriam Taboada Puga | 2 |
| Thiago Taffarel Rios Salazar | 1 |
| Susana Tojo Ramos | 1 |
| Marta Tome Gonzalez | 1 |
| Patricia Torres Álvarez | 1 |
| Angelica Trillo Trillo | 1 |
| Susana Uriz Prado | 1 |
| Luis Angel Varela Nuñez | 1 |
| Cristina Varga Martin | 1 |
| Jacobo Vazquez Marquez | 1 |
| Brais Vazquez Lamazares | 1 |
| Natalia Vazquez Iglesias | 1 |
| Andres Miguel Vazquez Bendaña | 1 |
| Vanesa Vazquez Modia | 1 |
| Begoña Vidal Maroño | 1 |
| Raquel Vidal Barreiro | 2 |
| Fatima Viñas Castro | 1 |
| Karina Vilas Cabeza | 1 |
| María Soledad Vilas Iglesias | 2 |
| Maria Villoch Garcia | 1 |
| Cristian Vispo Lopez | 1 |
| Sandra Viz Lasheras | 2 |
| Ouhao Zhu Huang | 2 |

**Affiliations:**

1.- Servizo Galego de Saude, Consellería de Sanidade, Galicia, Spain

2.- Genetics, Vaccines and Infectious Diseases Research Group (GENVIP), Instituto de Investigación Sanitaria de Santiago (IDIS), Santiago de Compostela, Galicia, Spain.
